# MRI Elastography Identifies Regions of Extracellular Matrix Reorganization Associated with Shorter Survival in Glioblastoma Patients

**DOI:** 10.1101/2022.11.07.22282021

**Authors:** Siri Fløgstad Svensson, Skarphéðinn Halldórsson, Anna Latysheva, Elies Fuster-Garcia, Trine Hjørnevik, Jorunn Fraser-Green, Robin A. B. Bugge, Jack Grinband, Sverre Holm, Ralph Sinkus, Einar O. Vik-Mo, Kyrre Eeg Emblem

**Affiliations:** Dept. of Physics and Computational Radiology, Division of Radiology and Nuclear Medicine, Oslo University Hospital, Oslo, Norway; Department of Physics, University of Oslo, Oslo, Norway; Vilhelm Magnus Laboratory, Dept. of Neurosurgery, Oslo University Hospital, Oslo, Norway; Department of Radiology, Division of Radiology and Nuclear Medicine, Oslo University Hospital, Oslo, Norway; BDSLab, Instituto Universitario de Tecnologías de la Información y Comunicaciones, Universitat Politècnica de València, València, Spain; The Intervention Center, Oslo University Hospital, Oslo, Norway; Department of Radiology, Columbia University, New York, USA; Department of Psychiatry, Columbia University, New York, USA; Division of Imaging Sciences and Biomedical Engineering, King’s College, London, United Kingdom; INSERM U1148, LVTS, University Paris Diderot, Paris, France

**Author notes:** Shared first-authorship (Siri Fløgstad Svensson, Skarphéðinn Halldórsson).

## Abstract

**Background:** The biomechanical tissue properties of glioblastoma tumors are heterogeneous, but the molecular mechanisms involved and the biological implications are poorly understood. Here, we combine magnetic resonance elastography (MRE) measurement of tissue stiffness with RNA sequencing of tissue biopsies to explore the molecular characteristics of the stiffness signal.

**Methods:** MRE was performed preoperatively in 13 patients with glioblastoma. Navigated biopsies were harvested during surgery and later classified as ‘stiff’ or ‘soft’ according to MRE stiffness measurements (|G*|_norm_). Twenty-two biopsies from eight patients were analysed by RNA sequencing.

**Results:** The mean whole-tumor stiffness was lower than in normal-appearing white matter. The surgeon’s biopsy stiffness evaluation did not correlate with the MRE measurements, which suggests that they measure different properties. Gene set enrichment analysis of the differentially expressed genes between ‘stiff’ and ‘soft’ biopsies showed that genes involved in extracellular matrix reorganization and cellular adhesion were overexpressed in ‘stiff’ biopsies. Supervised dimensionality reduction identified a gene expression signal separating ‘stiff ‘and ‘soft’ biopsies. Using the NIH Genomic Data Portal, 265 patients with glioblastoma were divided into patients with (n=63) and without (n=202) this gene expression signal. The median survival time of patients with tumors expressing the gene expression signal associated with ‘stiff’ biopsies was 100 days shorter than that of patients not expressing it (360 versus 460 days, hazard ratio: 1.45, P<0.05).

**Conclusion:** MRE imaging of glioblastoma can provide non-invasive information on intratumoral heterogeneity. Regions of extracellular matrix reorganization showed an expression signal correlated to shorter survival time in patients with glioblastoma.

**Importance of the study:** While the importance of biomechanical forces in glioblastoma is unquestioned, the underlying mechanisms are still not well understood, nor its clinical implications. Several methods exist to assess tissue stiffness, but MRE is unique in allowing measurements of stiffness *in vivo* and *in situ*. For the first time, we present molecular profiling of glioblastoma tissue correlated with *in situ* stiffness measurements. The transcriptomic profiles of ‘stiff’ and ‘soft’ biopsies showed that extracellular matrix reorganization was strongly associated with the ‘stiff’ biopsies, in particular collagen binding. Genes associated with innate immune processes were also upregulated in ‘stiff’ biopsies, indicating that these are active regions of the tumor. The association between gene expression in ‘stiff’ biopsies and survival is in concordance with previous reports of elevated extracellular matrix stiffness increasing glioblastoma aggression.

**Key Points:**

- MR Elastography can provide unique information on intratumoral heterogeneity preoperatively.
- MR Elastography identifies tumor regions of active extracellular reorganization
- Gene expression signal associated with increased stiffness negatively correlates with survival

## Introduction

Intratumoral heterogeneity is characteristic of glioblastoma (GBM) and is believed to be one of the key determinants of therapy failure^1^. Intratumoral heterogeneity stems from intrinsic genetic alterations as well as the inherent plasticity of GBM tumor cells that adapt to various microenvironmental factors^2,3^. One such factor is the biomechanical properties of the tumor and its microenvironment. The biomechanical properties of GBM affect how tumor cells interact with the local microenvironment and can contribute to tumor invasion^4,5^. Despite its importance intraoperatively and for tumor progression, little data exists on the physical characteristics and genetic determinants of biomechanical heterogeneity in GBM.

The biomechanical properties of the tissue within a GBM tumor can be highly variable, with subregions ranging from a soft and gel-like to a solid and dense consistency. These differences in biomechanical properties can impact the technical ease of resection and be an important determinant for operative planning. Resection of a stiff tumor that adheres to pia and vessels can result in damage to neighboring structures, while soft, liquescent tumors are more readily removed through gentle suction.

MR elastography (MRE) is an imaging technique that noninvasively measures the biomechanical properties of tissue. In contrast to intraoperative palpation by the surgeon, MRE provides a quantitative and objective measure of tissue stiffness, and characterizes its spatial distribution. Previous MRE studies in humans have found that GBM tumors differ from healthy brain in terms of shear stiffness and viscosity, and are spatially heterogeneous with respect to measured tissue stiffness^6,7^.

Here, we examine the intratumoral biomechanical heterogeneity of GBM tumors using preoperative MRE and MRI-localized biopsies. A comparison of high and low stiffness biopsies using RNA sequencing showed that genes involved in extracellular matrix organization were overexpressed in high stiffness biopsies and were a negative prognostic factor for patient survival. Our data demonstrates that MRE imaging of GBM provides unique information on tumor heterogeneity and helps identify probable regions of active extracellular matrix reorganization.

## Methods and Materials

This study was approved by the National Research Ethics Committee and the Institutional Review Board (2018/2464 and 2016/1791) and all patients gave written and informed consent. Thirteen patients (eight women and five men, median 56 years, range 38-75 years) with subsequent neuropathologically confirmed IDH wild-type GBM were prospectively included in the study. Four patients were excluded due to technically unsuccessful MRE and one patient was excluded due to MRI susceptibility artifacts caused by a cranium fixation item. Finally, one patient was excluded due to failed registration of biopsy coordinates.

### MR imaging

MRI exams were performed on a 3T clinical MRI scanner (Ingenia, Philips Medical Systems, Best, the Netherlands) using a 32-channel head coil. In addition to MRE, a clinical preoperative protocol was used, including a T1-weighted MPRAGE sequence (3D inversion recovery gradient echo, 1×1×1 mm^3^ resolution, 256×256×368 matrix, TR/TE=5.2/2.3 ms, shot interval 3000 ms, inversion delay 853 ms) acquired before and after administration of a gadolinium-based contrast agent, as well as T2-weighted (turbo spin echo, 0.6×0.6×4 mm^3^ resolution, 420×270×28 matrix, TR/TE=3000/80 ms) and T2-FLAIR sequences (turbo spin echo, 1×1×1 mm^3^ resolution, 252×249×183 matrix, TR/TE=4800/320 ms, inversion time 1650 ms). For the MRE, a gravitational transducer was attached to the side of the subject’s head to induce shear waves of 50 Hz in the brain^8^. Image acquisition was performed using a multi-shot gradient-echo MRE sequence, synchronized to the wave generator by a trigger signal^9^. Fifteen contiguous transversal slices were placed according to the tumor site and scanned using an isotropic resolution of 3.1 mm^3^, matrix size of 72 × 70, and FOV = 22 cm. Other scan parameters were: Flip angle = 20°, TR/TE = 384/12 msec, Cartesian readout, and a sensitivity encoding factor of 2. Hadamard motion encoding was performed using bipolar 13 mT/m motion-encoding gradients at 115 Hz in four directions^10^. Eight mechanical phase offsets were acquired throughout the period of the 25-Hz frequency component of the waveform. The actual mechanical vibration frequency was shifted to the second index of the Fourier transform, thus filtering out potential contributions from the 25, 75, and 100 Hz frequencies. The MRE acquisition time was 5.5 minutes, and was well tolerated across all patients.

### Surgery and tissue sampling

The surgery was performed by two neurosurgeons. In order to guide sampling, surgeons evaluated MRE data prior to surgery to plan for biopsies covering a range of MRE signal. During resection, 2-7 biopsies were taken from different parts of the tumor. All biopsies were situated in tumor or adjacent parts of the brain included in the planned resection prior to MRE evaluation, and covered both contrast-enhanced T1-weighted (CE-T1w) images and more diffusely infiltrating tumor demarcated by non-enhancing T1-weighted and pathological T2-FLAIR signal. Biopsies were taken early in the resection, prior to any major shift in the navigational accuracy. The biopsy locations were chosen according to varying stiffness as evaluated by the neurosurgeon. The surgeon evaluated tumor consistence according to a modified version of the grading scale from Zada et al. as either (1) softer than normal brain tissue, (2) similar in consistency to normal brain tissue, or (3) firmer than normal brain tissue^11^. Stereotactic guidance was provided by preoperative CE-T1w and T2-FLAIR images on a neuronavigation system (Brainlab Curve; Brainlab, Feldkirchen, Germany). The biopsies were snap-frozen immediately after extraction. The frozen biopsies were weighed, homogenized in a Tissuelyser (Qiagen, the Netherlands), and aliquoted for biomolecule extraction.

### Biopsy co-registration

The biopsy locations were recorded by screen captures of the neuronavigation interface at the time of tissue sampling and co-registered to the CE-T1w images using a semiautomated screen capture registration tool, allowing the determination of the Cartesian coordinates of each biopsy^12^. Next, a robust binary region-of-interest (ROI) was made for each coordinate by expanding one voxel in all three directions from the coordinate seed point. Finally, the positions of all ROIs were visually controlled by an experienced neuroradiologist. The same neuroradiologist also classified each biopsy as collected from either (1) contrast-enhancing tumor, (2) necrotic tissue, or (3) a nonenhancing region with pathological T2-FLAIR signal.

### MR Image Processing

From the MRE scan, maps of the magnitude of the shear storage modulus |G*| (tissue stiffness) and the shear phase angle ^φ^ (related to tissue viscosity, i.e. its ability to dissipate energy) were produced. Details about the MRE reconstruction can be found in Svensson et al.^7^. The volumetric CE-T1w images were co-registered to the MRE image space, using a nearest-neighbor interpolation in the nordicICE software (NordicNeuroLab AS, Bergen, Norway). The resulting transformations were applied to the binary ROI masks in CE-T1w space, resulting in a one-voxel seed point in MRE space. To make the analysis more robust to brain shift and co-registration issues, a ROI consisting of a trimmed mean of nine MRE voxels around the seed point was used, where the voxel with the highest and the lowest value were removed before averaging. **Figure 1A** shows an example of a biopsy location on a CE-T1w image and the corresponding |G*| map.

**Figure 1:**
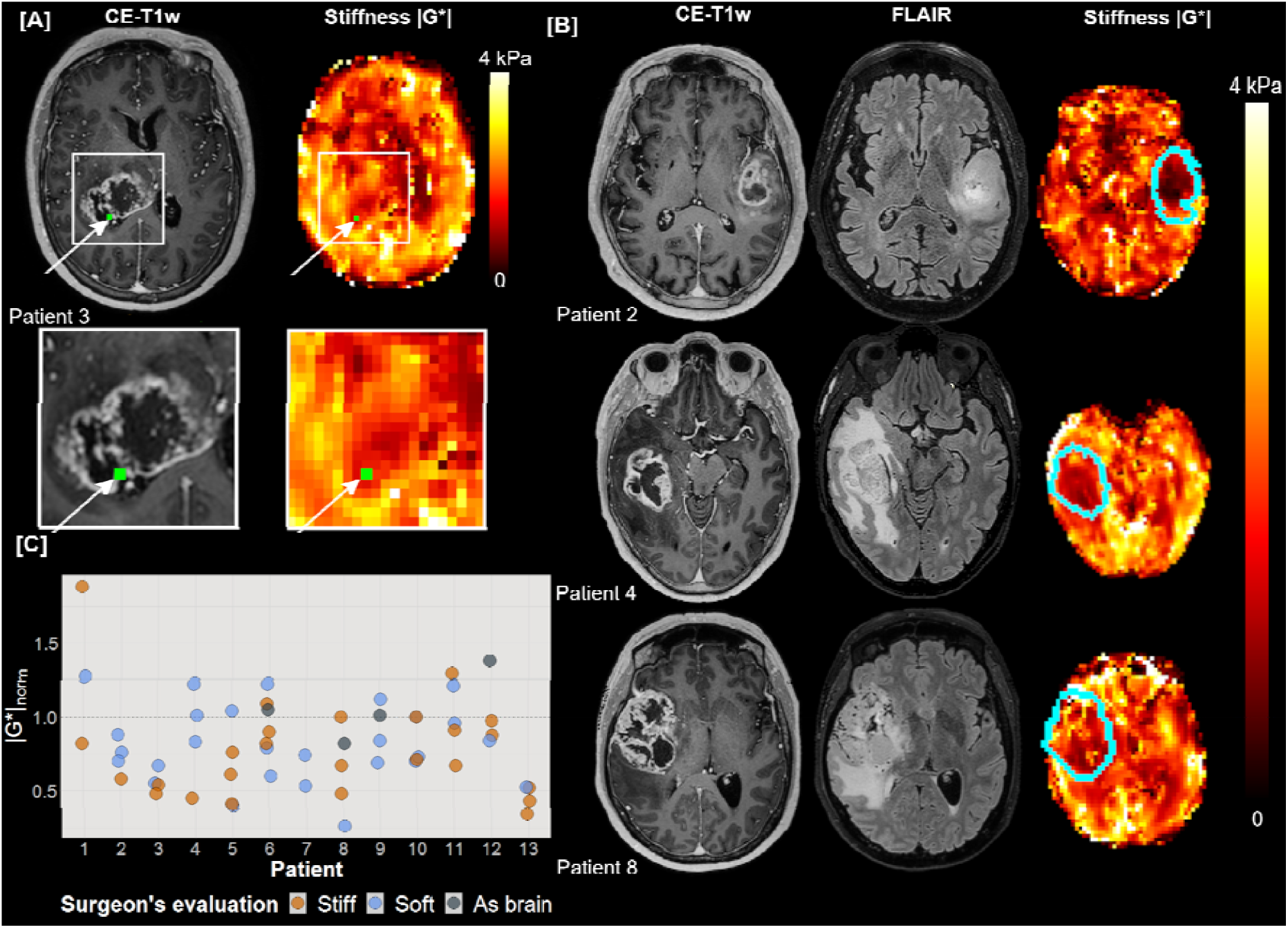
(A) Example of MRE imaging. The location of the tissue biopsy is shown in green in the contrast-enhanced T1-weighted image (CE-T1w) and the MRE stiffness map (|G*|). (B) Representative images for three patients. From left to right: CE-T1w images, T2-FLAIR images and |G*| map, with tumor outline shown in blue. (C) Stiffness measurements for all biopsies per patient (n=56). Normalized MRE-based stiffness |G*|_norm_ for all biopsies acquired for all patients (n=13). Biopsies evaluated by the surgeon as stiff are shown in orange (median = 0.71), biopsies evaluated as soft are shown in blue (median = 0.79) and biopsies found by the surgeon to be similar in consistency to healthy brain tissue are shown in gray (median = 1.03). The horizontal line at |G*|_norm_ = 1 shows the mean stiffness in each patient’s contralateral normal-appearing white matter.

Tissue segmentation was performed using Oncohabitats, a multiparametric system for glioblastoma heterogeneity assessment through MRI ^13^. This segmentation was performed for each patient based on pre- and post-contrast T1-weighted, T2-weighted and T2-FLAIR MRI scans, and resulted in segmentation of contrast-enhancing tumor, necrosis, peritumoral region of high signal on T2-FLAIR, and normal-appearing gray and white matter. The mean value of each patient’s contralateral normal-appearing white matter was used to normalize MRE measurements, resulting in |G*| ^14^.

### RNA Sequencing

For each patient, 2-4 biopsies were selected for RNA sequencing. Total RNA sequencing returned sequence counts for 22510 genes and other transcripts. The analyzed biopsies were classified as ‘stiff’ or ‘soft’ based on MRE, i.e., higher or lower |G*| _norm_ than the mean value of all biopsies within each tumor, respectively. Batch correction of RNA sequencing data was performed with the ComBat-seq package in R^15^. Normalization and differential expression of RNA sequencing data was done with the DESeq2 package in R^16^. The differential expression results were corrected for multiple testing using the Benjamini and Hochberg method, and adjusted p-value threshold was set to 0.05.

### Statistical analysis

Comparison of MRE measurements in tumor and normal-appearing tissue was performed using a Wilcoxon signed-rank test. MRE measurements and tumor volumes were compared using a Spearman’s rank order test. |G*|_norm_ measurements were compared to the surgeon’s evaluation using ordered logistic regression and to the radiological tissue type using multinomial regression. A significance level of P *=* 0.05 was assumed for all tests. Logistic regression was performed using Stata (version 17.0, StataCorp LLC, College Station, Texas, USA). Over-representation (OR) analysis and gene-set enrichment analysis (GSEA) were performed using the clusterProfiler package in R^17^. Principal component analysis (PCA), partial least-squares discriminant analysis (PLS-DA) and sparse PLS-DA were performed with the mixOmics package in R^18^.

### Survival analysis

Raw RNA sequencing reads from the The Cancer Genome Atlas (TCGA) and the Clinical Proteomic Tumor Analysis Consortium (CPTAC) projects were downloaded from the NIH Genomic Data Commons Data Portal along with sample metadata. Normal samples, control samples and duplicate samples were excluded, leaving 265 patient samples. Values were batch-corrected with ComBat-seq followed by normalization and rlog transformation in DESeq2. A sparse PLS-DA model containing 22 genes was trained using 22 patient samples, annotated as ‘stiff’ or ‘soft’, using the mixOmics package. Expression of these 22 genes was used to classify the external data using the ‘predict()’ function in the mixOmics package. Kaplan-Meier survival curves were produced with the survminer package in R. Cox regression analysis on survival data was performed in SPSS (version 28.0, IBM Corp, Armonk, NY).

### Code and Data Availability

The source code and data to reproduce all analyses and figures in this manuscript is available at https://github.com/SkabbiVML/stiffR.

## Results

The demographic data of the patient, tumor volumes, and mean |G*|_norm_ and φ _norm_ values are listed in **Supplementary Table 1**. Example MRE images are shown in **Fig. 1B**.

### GBM tumors are heterogeneous and softer than normal-appearing brain tissue

The mean stiffness |G*| was 20% lower in contrast-enhancing tumor than in normal-appearing white matter (P<0.001) and 30 % lower in necrosis than normal-appearing white matter (P<0.001). The mean shear phase angle φ was 10% lower in contrast-enhancing tumor than in normal-appearing white matter (P<0.001) and 8% lower in necrosis than in normal-appearing white matter (P<0.005). In the nonenhancing T2-FLAIR hyperintense regions, stiffness did not differ significantly from in normal-appearing white matter (P=0.8), but φ was 15% lower than in normal-appearing white matter (P<0.001). The median tumor volume (contrast-enhancing and necrotic regions combined) was 33 cm^3^ (range 7-78 cm^3^), and the median volume of the high signal region on T2-FLAIR was 45 cm^3^ (range 2-162 cm^3^). Mean |G*|_norm_ and φ _norm_ values did not correlate with the volume of tumor or edema.

The stiffness was heterogeneous both between and within tumors (**Fig. 1C**). Some patients (e.g., patient 13) had |G*|_norm_ <1 for all biopsies, while others displayed higher stiffness values for some biopsies (e.g., patient 1). The median ratio between the biopsy with highest and lowest stiffness of a patient was 1.6 (range 1.4-3.9). **Table 1** shows |G*|_norm_ and the surgeon’s evaluation of biopsies that were analyzed by RNA sequencing. The table also shows whether the biopsy was taken from contrast-enhancing tumor, necrosis or a region with high signal on T2-FLAIR. Measured stiffness |G*|_norm_ was not correlated to the radiological tissue type of the biopsy (P=0.06).

**Table 1.**
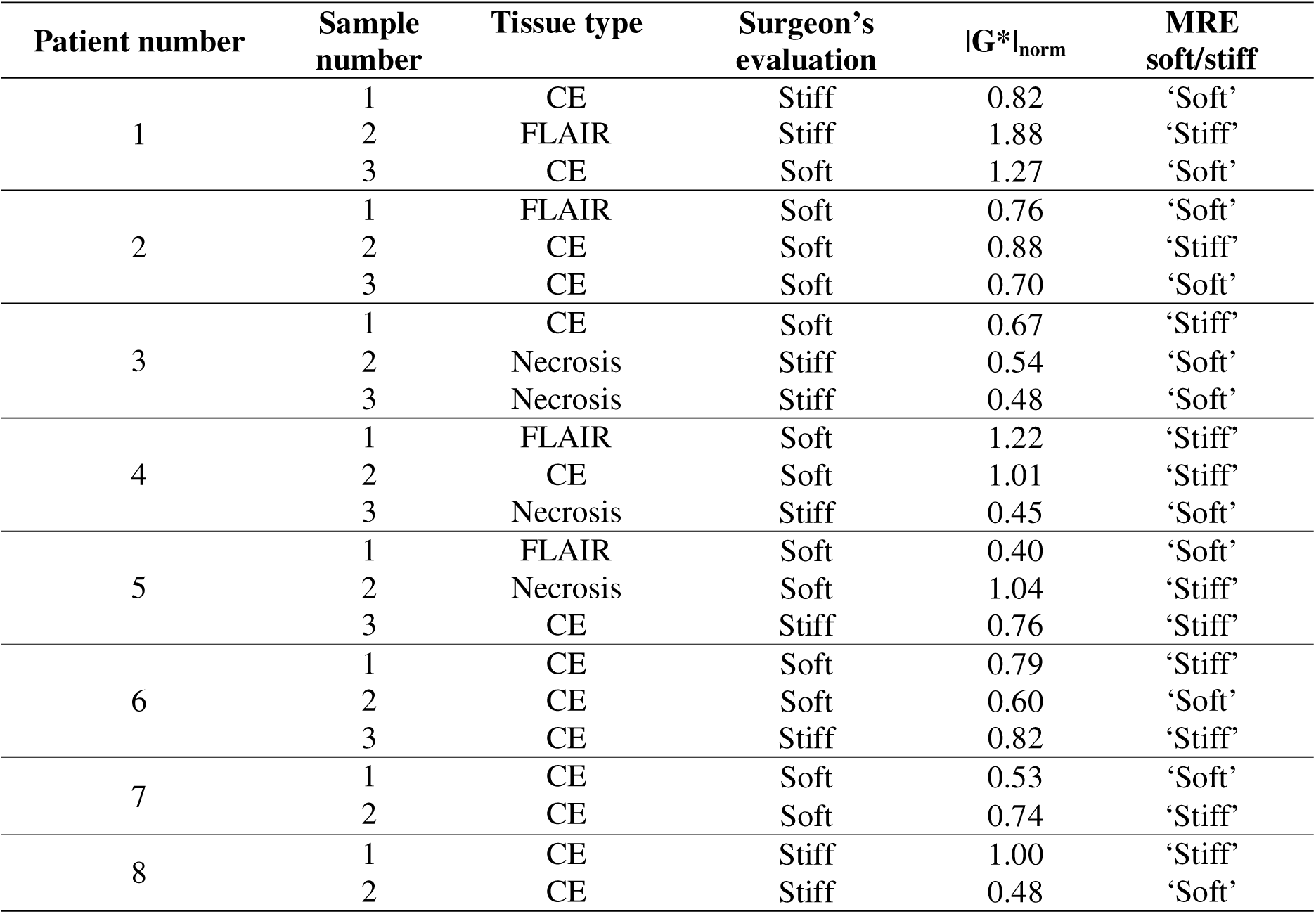
Biopsies used in RNA sequencing. The tissue type of each biopsy point was radiologically evaluated, using CE-T1w and T2-FLAIR images, as contrast-enhancing tumor (CE), region with high signal on T2-FLAIR (FLAIR) or necrosis. Surgeon’s evaluation of the consistency of each biopsy (stiff or soft compared to normal brain parenchyma), |G*|_norm_, and MRE biopsy classification (‘stiff’ or ‘soft’, i.e. higher or lower |G*|_norm_ than the mean value of all biopsies within each tumor, respectively).

The surgeon’s evaluation of biopsy stiffness during surgery and |G*|_norm_ did not correlate (P=0.58), suggesting that the two entities are independent measures.

### Gene expression associated with GBM stiffness

To evaluate the molecular differences between stiff and soft tissue biopsies, we performed total RNA sequencing on 22 biopsies from 8 GBM tumors (patients 1-8). Unsupervised dimensionality reduction by principal component analysis (**Supplementary Fig. 1A**) did not identify tissue stiffness as measured by MRE as a strong source of variance within the data. However, supervised dimensionality-reduction with partial least-squares discriminant analysis (PLS-DA) identified an expression signal that separated ‘stiff’ from ‘soft’ biopsies within each tumor. Tuning of the PLS-DA parameters (5-fold cross-validation, 100 repeats) indicated that a minimal sparse PLS-DA model containing 22 genes was sufficient to separate 22 patient samples based on the measured tissue stiffness (**Supplementary Fig. 1C**).

Differential gene expression analysis between ‘stiff’ and the ‘soft’ biopsies per patient found that 196 genes were differentially expressed based on an adjusted p-value of 0.05 (**Fig. 2A, Supplementary Table 2**). Of these, 122 were upregulated in ‘stiff’ biopsies, while 74 were upregulated in ‘soft’ biopsies. Normalized expression levels of differentially expressed genes in every biopsy show that ‘stiff’ or ‘soft’ biopsies tend to cluster together, and biopsies within individual patients also show similar expression profiles (**Fig. 2B**). Due to the limited size of the dataset, differential expression may depend on samples from a single patient. Therefore, to explore the robustness of the differential expression, we performed sequential differential expression analysis, leaving out all samples from a single patient in each iteration. Patient-wise leave-one-out cross-validation identified a set of 43 genes (35 in ‘stiff’ biopsies and 8 in ‘soft’ biopsies) that were found to be differentially expressed on every iteration (**Table 2**).

**Table 2.**
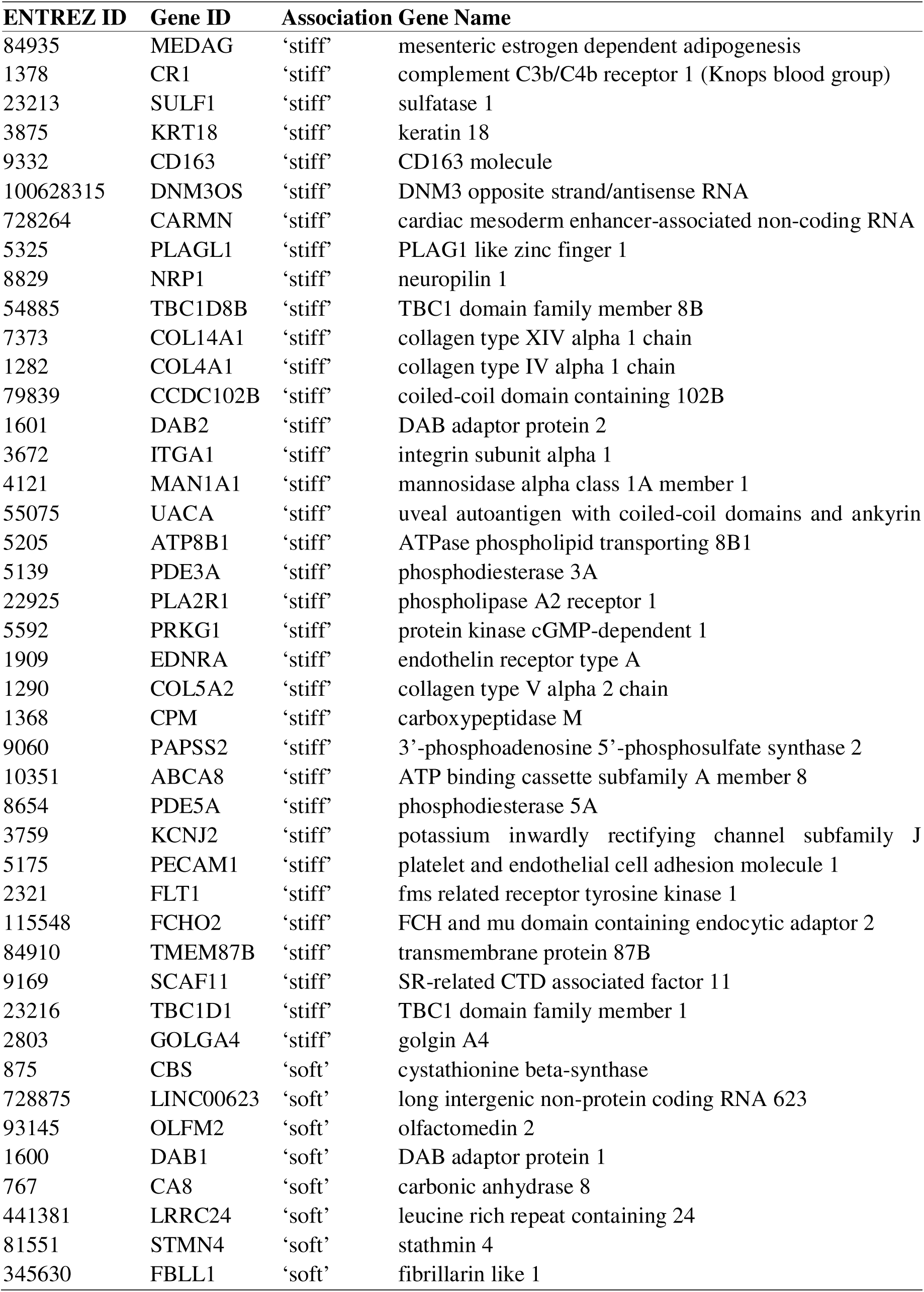
Differentially expressed genes: List of most stable differentially expressed genes between ‘stiff’ and ‘soft’ biopsies after patient-wise leave-one-out validation.

**Figure 2.**
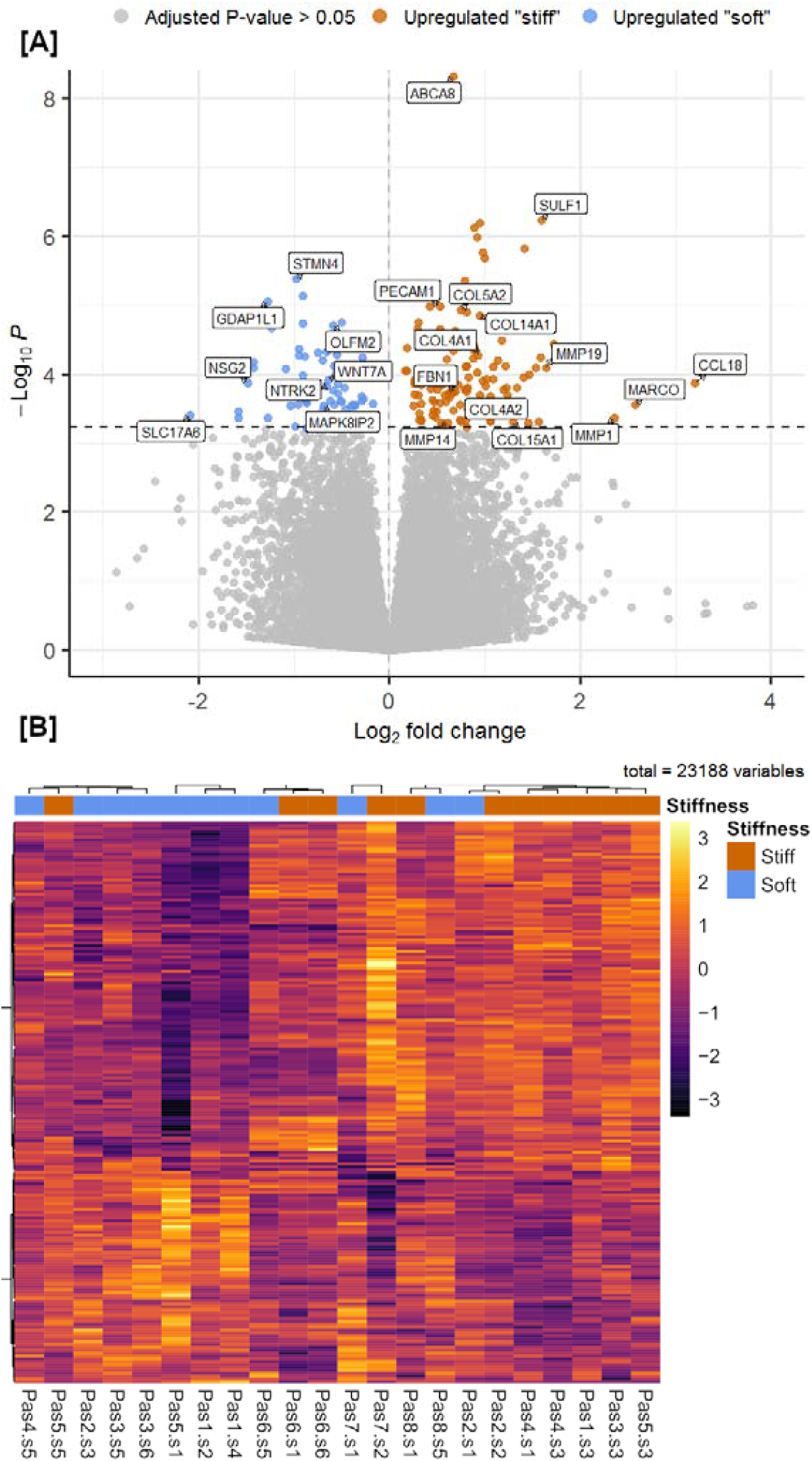
Differential gene expression between ‘stiff’ and ‘soft’ biopsies in GBM tumors. (A) Volcano plot summarizing the results of differential gene expression between ‘stiff’ and ‘soft’ biopsies. The magnitude of change in expression between ‘stiff’ and ‘soft’ biopsies is shown along the x-axis, and the statistical significance is shown along the y-axis. 521 genes were found to have differential expression between the groups with an adjusted p-value < 0.1, 196 genes with an adjusted p-value < 0.05 (shown in blue and red). (B) Heatmap of differentially expressed genes, where biopsies are grouped according to the pattern of gene expression. All genes that passed an adjusted p-value threshold of 0.05 (196 genes) are included in the heat map, along the y-axis. The analyzed biopsies are shown along the x-axis. The color gradient shows the changes of gene expression, the expression of genes is scaled across all biopsies. “Pas” represents patient number, “s” represents biopsy number.

Classifying biopsies into stiff or soft by the surgeon’s evaluation was also performed, but no significant difference in gene expression was found between biopsies classified in this way.

### Functional annotation of stiffness-associated gene expression

To evaluate the structural and functional importance of differentially expressed genes, we performed a gene set enrichment analysis (GSEA) of genes associated with increased biopsy stiffness using the Gene Ontology (GO) and Reactome databases^19,20^. The GO-terms with the highest association to ‘stiff’ biopsies represent components of the extracellular matrix, cellular adhesion and innate immunity (**Fig. 3A**). Similarly, among the most significantly enriched Reactome pathways associated with ‘stiff’ biopsies were extracellular matrix organization, integrin cell surface interactions, and neutrophil degranulation (**Supplementary Fig. 2)**. In contrast, GO-terms and Reactome pathways with highest association with ‘soft’ biopsies largely represented normal neuronal functions such as regulation of membrane potential and neurotransmitter receptor complex although associations to DNA methylation and rRNA regulation were also found. Pathway enrichment maps show three distinct clusters of GO-terms with varying degrees of overlap. GO-terms associated with extracellular matrix reconstruction are upregulated in stiff samples. There is some overlap of genes associated with extracellular matrix terms and terms associated with effector cells of the innate immune system (neutrophils and granulocytes) which are also upregulated in stiff samples **(Fig. 3B)**. A third cluster of pathways, representing neuronal synapses and synaptic membranes, is upregulated in ‘soft’ biopsies.

**Figure 3.**
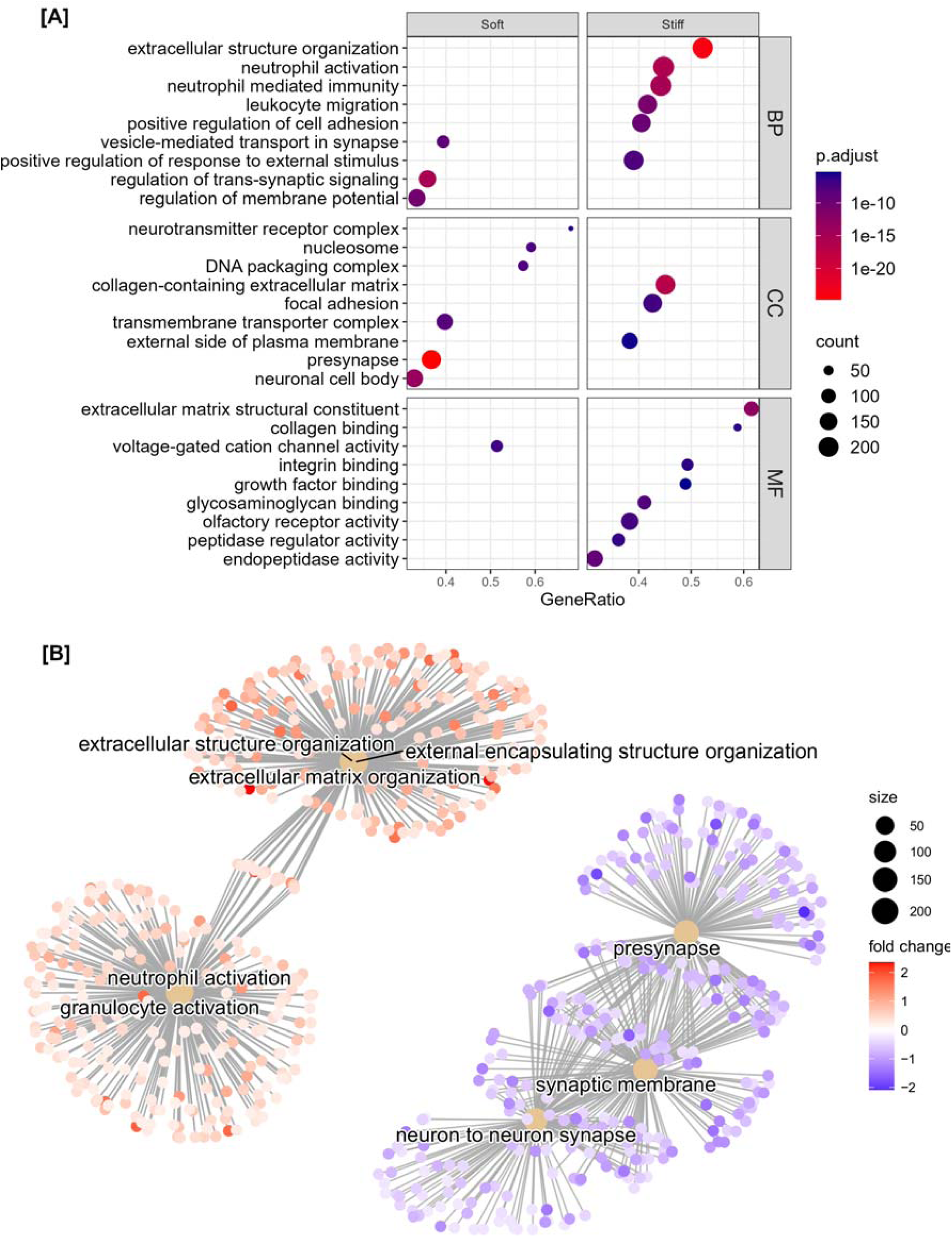
Gene-set enrichment analysis of differentially expressed genes in ‘soft’ and ‘stiff’ tumor biopsies. (A) Dot plot representing the terms most highly enriched in each Gene Ontology category. BP = Biological process, CC = cellular component, MF = molecular function, p.adjust = p-value adjusted for multiple testing. (B) Pathway enrichment map of the GSEA results. Central nodes represent Gene Ontology terms, colored dots represent differentially expressed genes (red = upregulated in ‘stiff’ biopsies, blue = upregulated in ‘soft’ biopsies).

Over-representation analysis of the 196 genes found to be upregulated in ‘stiff’ or ‘soft’ biopsies (adjusted P-value < 0.05, **Supplementary Table 2)** was largely concordant with the results from the GSEA: ‘Stiff’ biopsies were associated with collagen-containing matrix reorganization, focal adhesion, and immune cell activation/migration, while ‘soft’ biopsies were associated with normal synaptic activity and, to a lesser extent, DNA packaging and nucleosomes. Over-representation analysis of Reactome pathways identified ‘Extracellular matrix organization’ as strongly associated with ‘stiff’ biopsies while DNA methylation and RNA polymerase I promoter opening were associated with ‘soft’ biopsies (**Supplementary Fig. 3)**.

### Tissue stiffness is a negative prognostic factor for patient survival

Extracellular matrix reorganization and increased tissue stiffness have been associated with tumor cell infiltration in glioma^5^. Based on our findings, we hypothesized that stiffer tumor regions could represent regions of the GBM important for tumor progression and hence patient survival. To further study the effect of the gene expression signal that distinguished our ‘stiff’ and ‘soft’ biopsies, we evaluated RNA transcription profiles of 265 GBM tumors from two studies available in the NIH Genomic Data Commons Data Portal (168 biopsies from TCGA and 97 biopsies from CPTAC)^21^. Expression patterns of the 22 stable genes selected by PLS-DA were used to classify the tumors *with* this gene expression signal (n = 63) and tumors *without* it (n = 202). Survival analysis showed that the median survival time of patients carrying tumors expressing this gene signal was 100 days shorter than that of patients without this gene expression signal, from a median of 460 to 360 days (**Fig. 4**). Cox regression analysis showed that this gene expression signal had a significant impact on survival, with a 45% higher risk of death at any given time for patients with this gene expression signal. This result was significant after adjusting for age, gender, and type of treatment (hazard ratio: 1.45, 95% confidence interval: 1.043-2.015, P<0.05).

**Figure 4:**
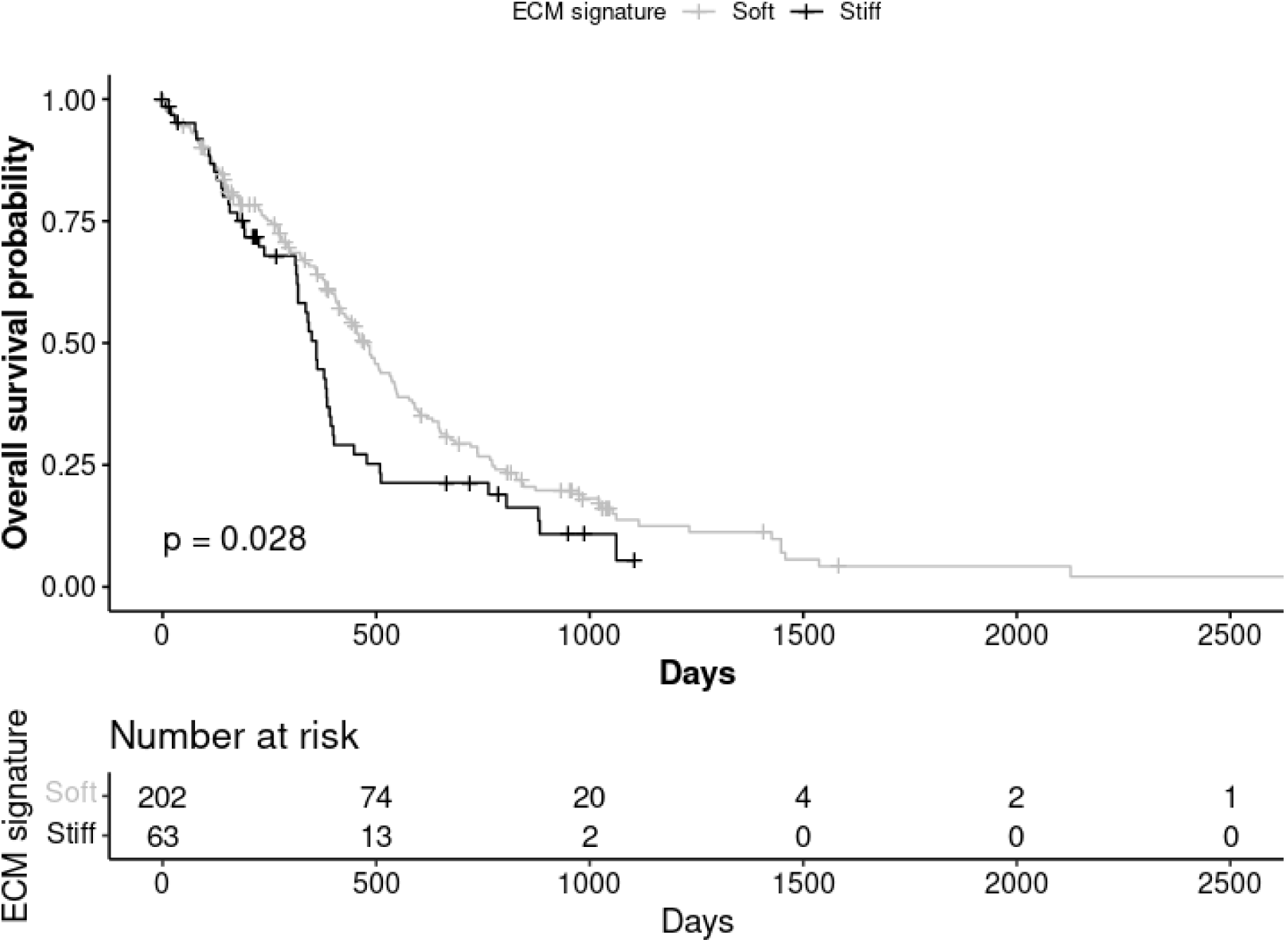
Survival analysis using data from TCGA and CPTAC. The median survival time of patients carrying tumors expressing the 22 genes associated with ‘stiff’ biopsies (n = 63, shown in black) was 100 days shorter than that of patients who did not express these genes (n = 202, shown in gray).

## Discussion

In our study, we compared the transcriptomic profiles of stiff and soft GBM tissue biopsies as measured by MRE. We found that extracellular matrix reorganization, focal adhesion, and neutrophile-mediated immune responses were associated with increased stiffness within the tumor. Our quantitative measure of stiffness in each biopsy location did not correlate with the surgeon’s subjective evaluation based on palpation. The ‘stiff’ and ‘soft’ biopsies as quantified by MRE could be separated by a gene expression signal of 22 genes. Finally, we showed that the expression signal found in ‘stiff’ biopsies is associated with decreased survival in patients with GBM.

Previous MRE studies on GBM tumors have reported decreased mean tumor stiffness of GBM compared to radiographically normal-appearing white matter^6^. This is in accordance with our findings that tumor tissue is on average softer than contralateral white matter. However, the stiffness within each tumor was also highly variable with regions of both high and low values compared to normal-appearing tissue.

The level of tissue stiffness in and around a glioblastoma *in vivo* is not well understood. At the single-cell level, glioma cells have been measured to be stiffer than normal brain cells^22,23^. On the nanoscale level, several studies using atomic force microscopy (AFM) report that GBM tissue is stiffer than non-tumor tissue^24-26^. However, GBM has also been reported to be softer than the normal brain using AFM in animal models^27^. Additionally, several *in vitro* studies have found GBM cells to be softer than normal fibroblasts and grade III glioma cells^28,29^. Ciesluk *et al*. found GBM tumor stiffness, as measured by AFM, to be higher on average than healthy tissue but also highly heterogeneous^26^. These findings on a microscale level are in contrast to previous bulk measurements obtained by MRE^6^. Hence, the measured stiffness in GBM varies both with the method and on the scale at which it is measured^26,30^. In contrast to these *in vitro* methods, MRE uniquely measures tissue stiffness *in vivo* and *in situ*.

A major source of tissue stiffening in GBM is believed to be restructuring of the extracellular matrix although definitive evidence of this is yet to be produced^24^. To the best of our knowledge, this is the first time molecular profiling of GBM tissue has been correlated with *in situ* MRE stiffness measurements. The transcriptomic profiles of ‘stiff’ and ‘soft’ biopsies in our study showed that extracellular matrix reorganization was strongly associated with ‘stiff’ biopsies, in particular collagen-related processes. Levels of fibrillar collagens in the healthy brain are low compared to the rest of the body, but in glioma, collagen levels are elevated and play a vital role in driving tumor progression^31^. Gene sets associated with innate immune processes, such as neutrophil activation, were also upregulated in ‘stiff’ biopsies, indicating that these are active regions of the tumor^32^. Thus, our findings support the idea that as the tumor progresses, it remodels its environment, producing a stiffening of the extracellular matrix. Elevated extracellular matrix stiffness has been shown to increase GBM aggression^33^ and increase proliferation^5,34^. Several of the genes found in our study to be upregulated in ‘stiff’ biopsies have previously been shown to play a role in glioma malignancy. *NRP1* and *DAB2* have been linked to glioma progression^35,36^, *PECAM1* correlates with GBM aggressiveness^37^, *CD163* is positively associated with the glioma malignancy grade^38^, and *Flt1* promotes invasion and migration of glioblastoma cells^39^. *CR1, PLAGL1, COL4A1*, and *COL5A2* have all been shown to correlate with shorter survival^40-43^.

When our data was compared to previously published transcriptomic profiles of GBM samples, we found that median survival was significantly shorter in patients with tumors that exhibited the gene expression signal associated with ‘stiff’ biopsies. This indicates that the genetic and molecular processes we detect in stiff tumor biopsies play a role in the malignant progression of tumors. It should be noted that while our data consists of multiple biopsies per patient with MRE stiffness evaluations, only one tissue biopsy per patient was available in the external data sets. We assume that this tissue biopsy is representative of the entire tumor. We do not claim that the tumors in these datasets are stiff or soft, rather, we examined the prognostic relevance of stiff or soft expression signals for GBM patients. However, a significant correlation between the proportion of highly stiff ECM areas within a GBM tumor and worse patient prognosis has previously been shown, suggesting that elevated ECM stiffness can foster GBM aggression^33^.

In contrast to AFM, MRE provides a stiffness map of the entire tumor and the surrounding tissue, and is therefore uniquely able to capture the heterogeneity of the biomechanical properties of a tumor prior to surgery. Previous work comparing the evaluation of tumor stiffness by neurosurgeons with MRE in meningiomas and in pituitary adenomas found that the measured stiffness correlated positively with the surgeon’s evaluation^44,45^. However, in these studies, the mean stiffness value for the entire tumor was reported. From tumor to tumor, meningiomas are known to vary in stiffness, from very firm to very soft^44^. In patients with gliomas, the surgeons’ haptic impression has been found to vary widely and therefore was not suitable as a gold standard of tumor consistency^46^. This illustrates the challenge of comparing MRE measurements with the surgeon’s impression, especially for small regions of interest. MRE probes the shear properties of tissue, while the probing by surgical tools involves a more complex process, as the tissue could also be compressed and compromised. Furthermore, tumor growth can compress surrounding tissue, generating solid stress due to swelling^47,48^. Several studies have found that MRE is sensitive to compressive stress^49,50^. Opening the skull during a craniotomy changes the pressure conditions in the brain, which may affect the perceived tissue stiffness compared to the MRE measurements, performed while the skull was still intact. When classifying biopsies using the surgeon’s evaluation rather than MRE, no significant difference in gene expression was found between biopsies evaluated as stiff and soft.

In conclusion, MRE identifies regions of malignant extracellular matrix reorganization with an expression signal correlated to shorter survival time in patients with glioblastoma. Thus, MRE may be a powerful tool for characterizing tumor heterogeneity during pre-surgical planning.

## Data Availability

All data produced in the present study are available upon reasonable request to the authors.

