## Supplementary material for "MRI Elastography Identifies Regions of Extracellular Matrix Reorganization Associated with Shorter Survival in Glioblastoma Patients"

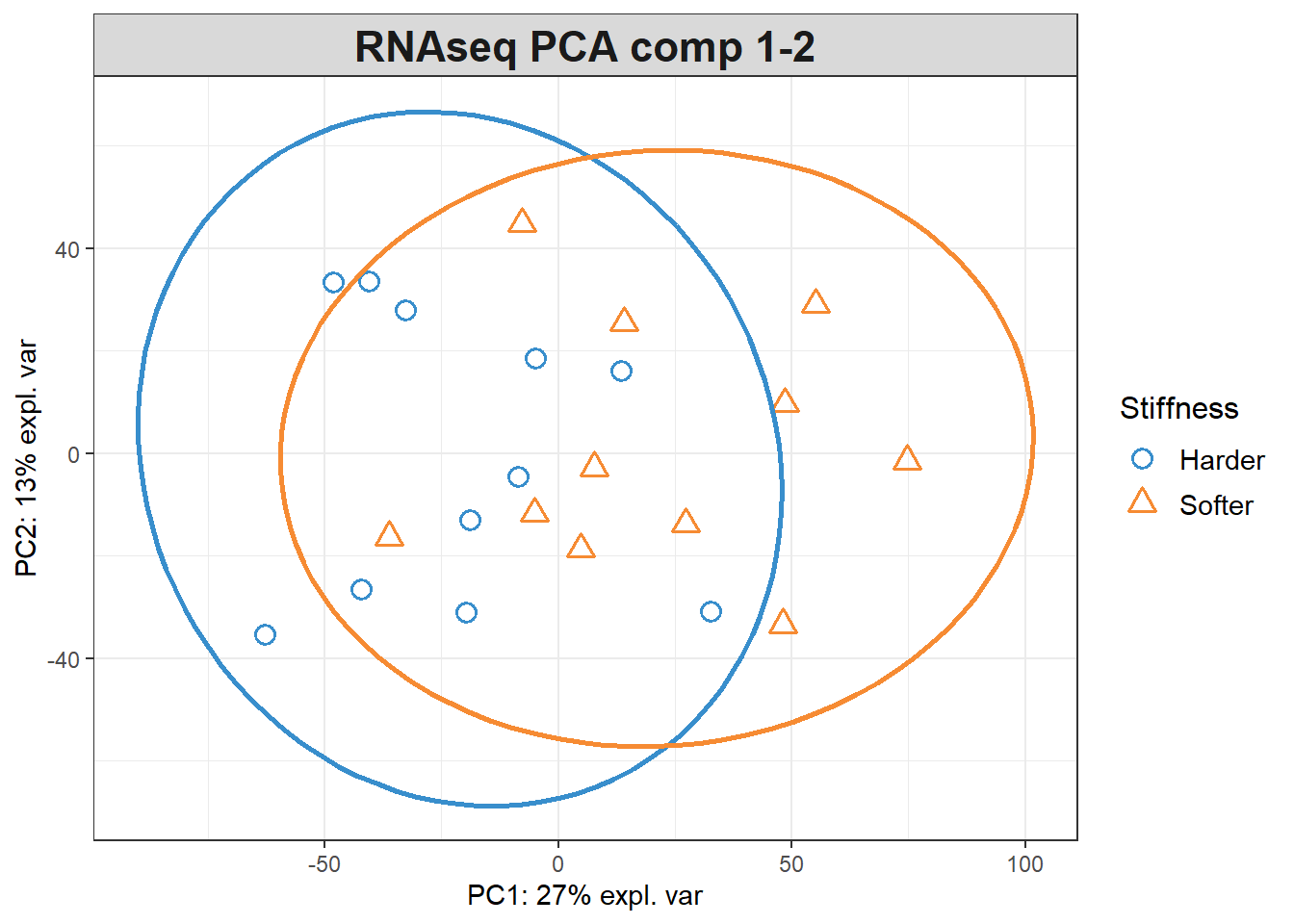

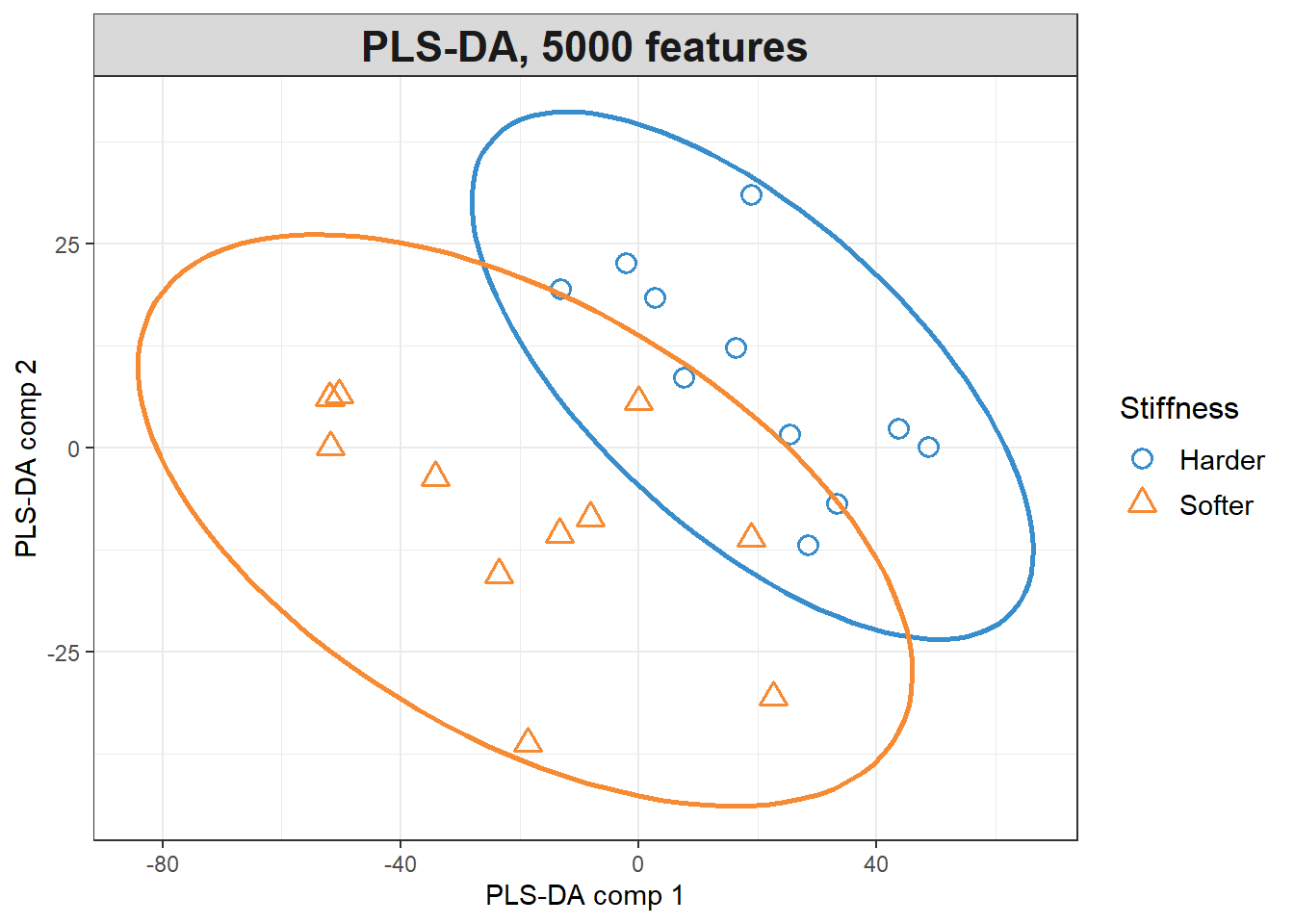

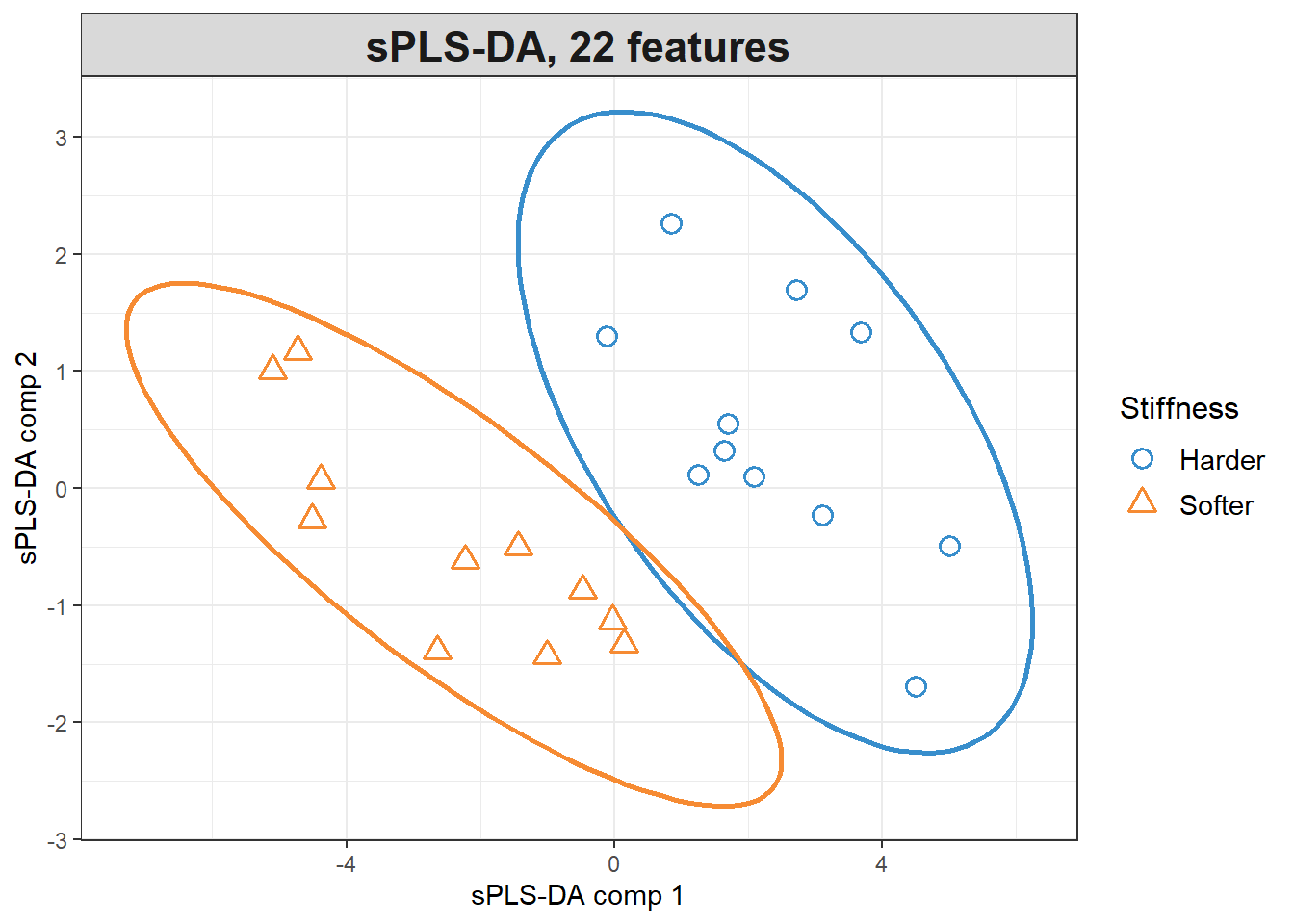

***Supplementary Figure 1:* *Clustering of glioblastoma biopsies based on |G*|_norm_.***  *Biopsies from each patient were classified as ‘stiff’ (blue circles) or ’soft’ (orange triangles) based on the mean stiffness of all biopsies from the same patient. A) Multilevel principal component analysis, accounting for patient variability, shows weak separation of ‘stiff’ and ‘soft’ biopsies along the first principal component. B) Partial least squares differential analysis (PLS-DA) based on the 5000 RNA transcripts with highest variance in the data shows that an expression signal can be found along the first component that distinguishes ‘stiff’ biopsies from ‘soft’ biopsies. C) The tuning of the PLS-DA results revealed that a set of 20 genes along the first component and 2 genes along the second component was sufficient to completely separate the ‘stiff’ and ‘soft’ biopsies.*

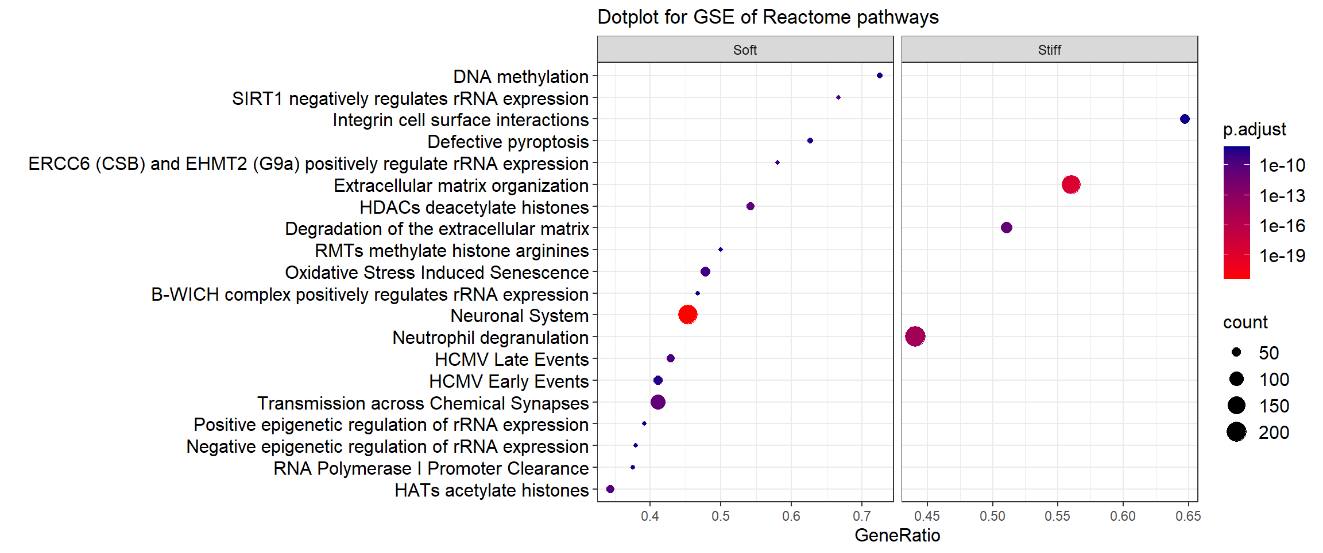

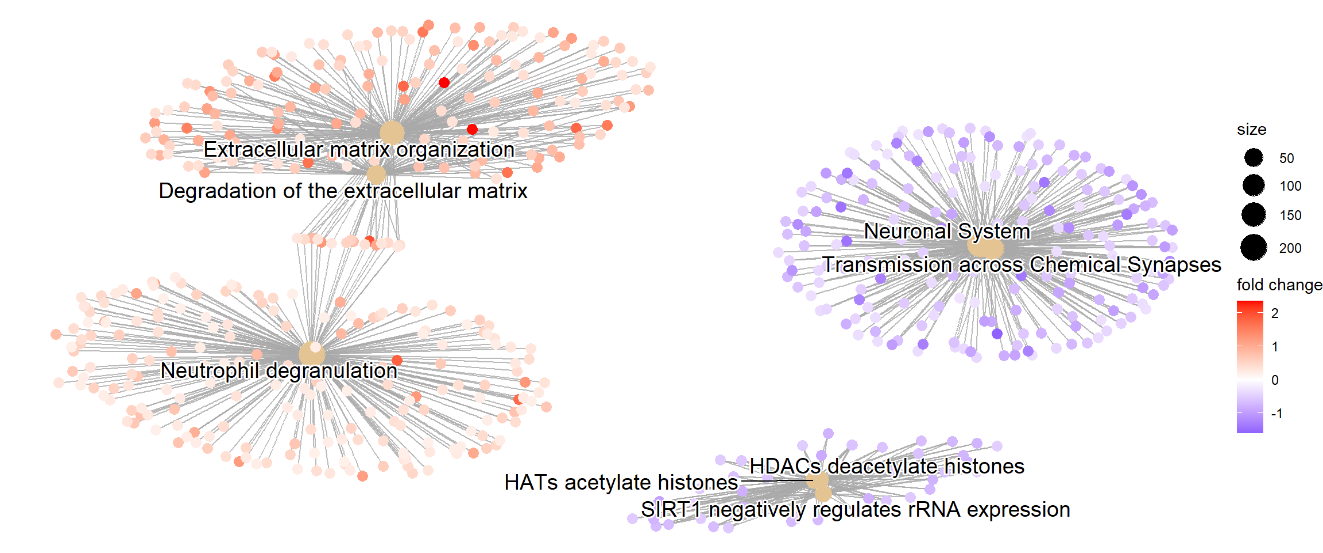

***Supplementary Figure 2. Gene-set enrichment analysis (Reactome pathways) of differentially expressed genes in ‘soft’ and ‘stiff’ tumor biopsies.*** *A) Dotplot representing the terms most highly enriched in Reactome pathways, p.adjust = p-value adjusted for multiple testing. B) Pathway enrichment map of the GSEA results. Central nodes represent Reactome pathways, colored dots represent differentially expressed genes (red = upregulated in ‘stiff’ biopsies, blue = upregulated in ‘soft’ biopsies).*

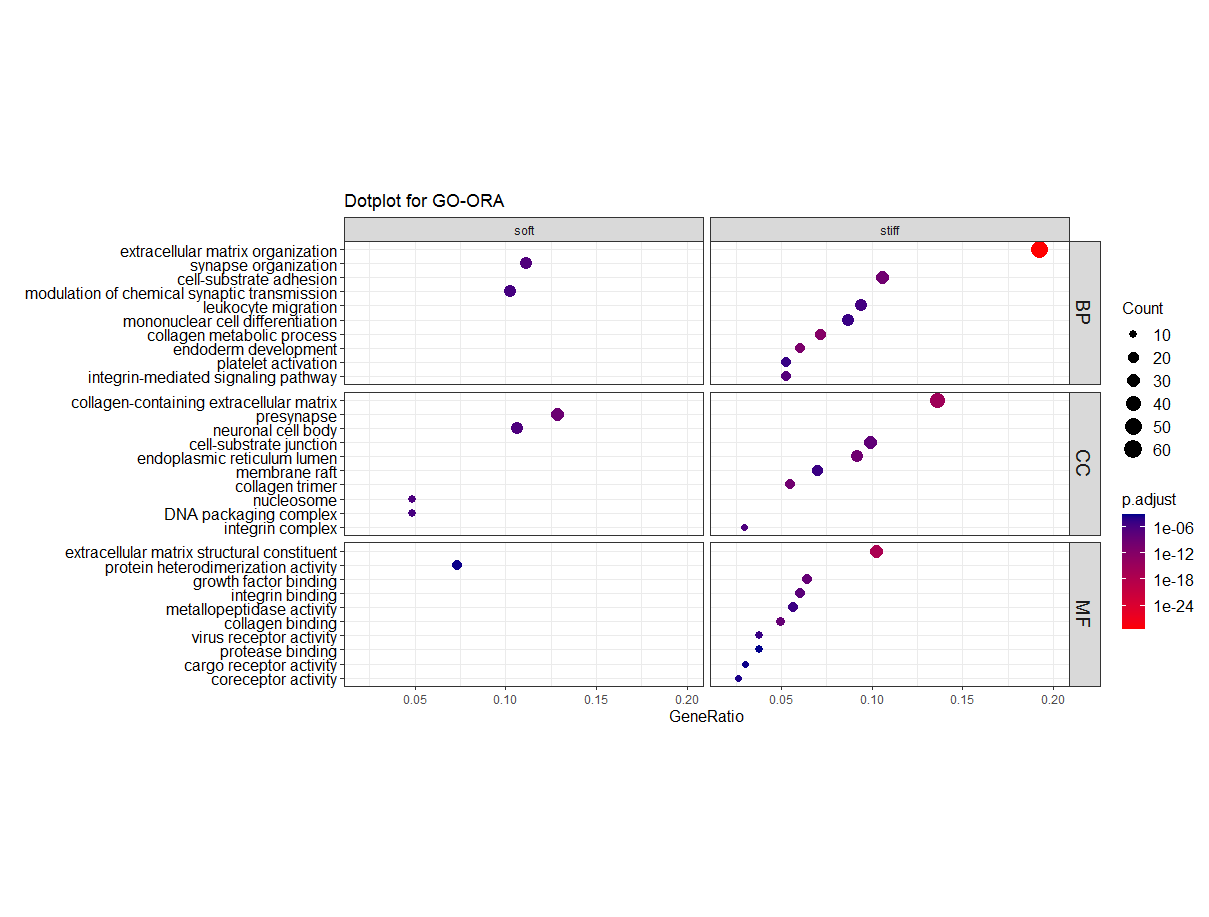

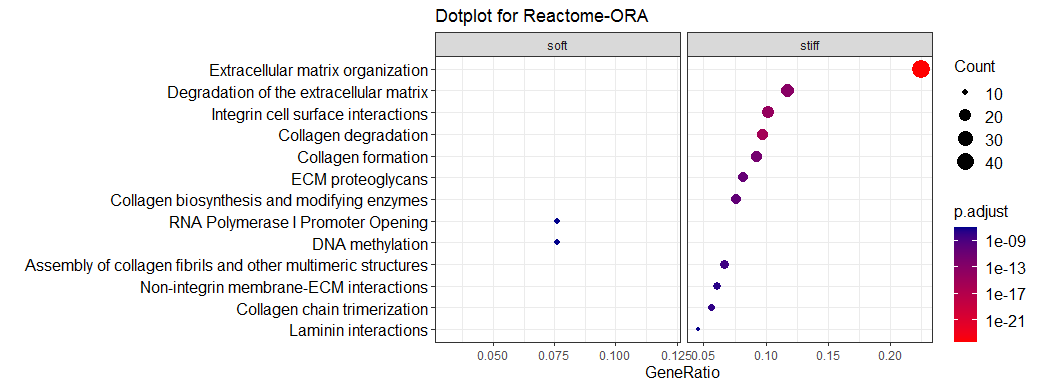

***Supplementary Figure 3:* *GO and Reactome Over-representation analysis.*** *A) Top 10 GO gene sets in each category (BP = biological process, CC = Cellular component, MF = Molecular function) with lowest adjusted p-value in ‘stiff’ and ‘soft’ biopsies.* *B) Top 13 Reactome pathways with lowest adjusted p-value in ‘stiff’ and ’soft’ biopsies.*

To evaluate the cellular and microenvironmental signatures within biopsies based on gene expression profiles, we performed a cell type enrichment analysis using the xCell signature set ^1^. Although we found a high variability in cellular composition as predicted by xCell between the biopsies, the stiffer biopsies tended towards higher astrocyte composition, while the ‘soft biopsies’ showed a higher score for neurons. Furthermore, a higher aggregated microenvironment score was seen in the ‘stiff’ biopsies **(Supplementary Fig. 4).**

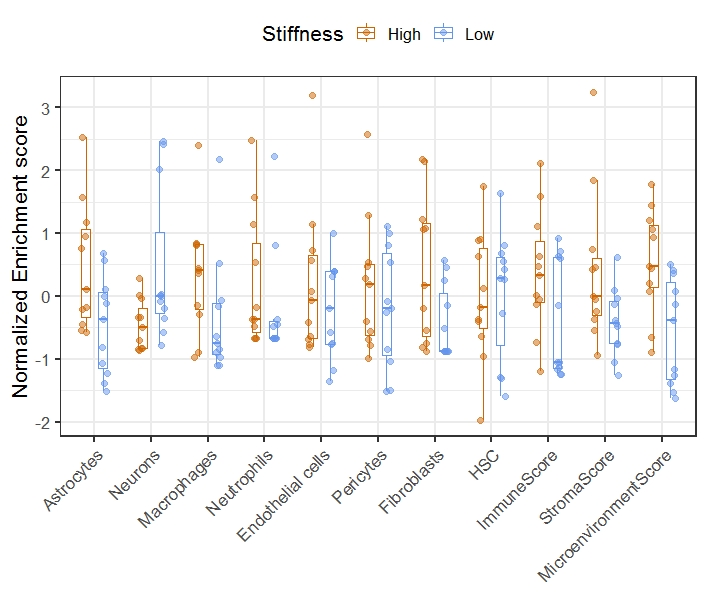

***Supplementary Figure 4:*** ***Cell-type enrichment in bulk RNA sequencing with xCell. ‘****Stiff’ biopsies shown in blue, ‘soft’ biopsies shown in orange.*

| **Patient age** | **Patient gender** | **Tumor location** | **Volume CE tumor [cm^3^]** | **Volume necrosis [cm^3^]** | **Volume FLAIR signal [cm^3^]** | **Mean \|G*\|_norm_ CE tumor** | **Mean \|G*\|_norm_ necrosis** | **Mean \|G*\|_norm_ FLAIR signal** | **Mean** φ**_norm_ CE tumor** | **Mean** φ**_norm_ necrosis** | **Mean** φ**_norm_ FLAIR signal** |
| --- | --- | --- | --- | --- | --- | --- | --- | --- | --- | --- | --- |
| 45-49 | Male | Parietal | 7 | 4 | 51 | 0.80 | 0.66 | 0.99 | 0.99 | 1.01 | 0.95 |
| 75-79 | Male | Temporal | 21 | 12 | 40 | 0.78 | 0.48 | 1.08 | 0.75 | 0.79 | 0.91 |
| 60-64 | Female | Basal ganglia | 29 | 13 | 43 | 0.73 | 0.69 | 0.83 | 0.82 | 0.97 | 0.83 |
| 55-59 | Male | Temporal | 28 | 11 | 162 | 0.80 | 0.64 | 0.89 | 0.79 | 0.67 | 0.82 |
| 35-39 | Female | Parietal | 14 | 4 | 92 | 0.74 | 0.76 | 1.05 | 0.71 | 0.54 | 0.80 |
| 60-64 | Male | Temporal | 28 | 19 | 39 | 0.90 | 0.98 | 1.01 | 0.81 | 0.71 | 0.83 |
| 50-54 | Male | Temporal | 7 | 3 | 45 | 0.84 | 0.65 | 1.21 | 0.97 | 0.98 | 0.80 |
| 50-54 | Female | Temporal | 49 | 25 | 61 | 0.80 | 0.80 | 0.95 | 0.97 | 0.92 | 0.86 |
| 40-44 | Female | Temporal | 5 | 2 | 50 | 0.81 | 0.85 | 1.09 | 0.75 | 0.71 | 0.93 |
| 65-69 | Female | Parietal | 11 | 8 | 2 | 0.98 | 0.77 | 1.21 | 0.88 | 0.82 | 0.85 |
| 65-69 | Female | Parieto-occipital | 39 | 10 | 42 | 0.93 | 0.70 | 0.99 | 0.88 | 1.00 | 0.95 |
| 60-64 | Female | Parieto-occipital | 33 | 45 | 35 | 0.81 | 0.75 | 0.94 | 0.78 | 1.26 | 0.87 |
| 45-49 | Female | Parietal | 13 | 13 | 52 | 0.61 | 0.56 | 0.92 | 0.76 | 0.97 | 0.76 |

**Supplementary Table 1: *Patient characteristics.*** *Patient age group, tumor location, volume, and MRE measurements (normalized to each patient’s contralateral normal-appearing white matter*) *in contrast-enhancing tumor, necrosis, and region with high signal on T2-FLAIR. The first eight patients were used in RNA sequencing analysis.*

| **ENREZ_ID** | **Symbol** | **baseMean** | **log2Fold Change** | **lfcSE** | **P-value** | **Adju. P-value** |
| --- | --- | --- | --- | --- | --- | --- |
| 10351 | ABCA8 | 1204.883261 | 0.671257 | 0.114681 | 4.82E-09 | 7.70E-05 |
| 23213 | SULF1 | 2151.787754 | 1.595155 | 0.319137 | 5.78E-07 | 0.003052 |
| 54885 | TBC1D8B | 118.289721 | 0.950754 | 0.190976 | 6.41E-07 | 0.003052 |
| 4121 | MAN1A1 | 903.7229709 | 0.890446 | 0.180097 | 7.64E-07 | 0.003052 |
| 79839 | CCDC102B | 1032.172753 | 0.922098 | 0.188653 | 1.02E-06 | 0.003258 |
| 3875 | KRT18 | 62.60869999 | 1.415651 | 0.294371 | 1.52E-06 | 0.003951 |
| 8829 | NRP1 | 4671.780854 | 0.982309 | 0.205396 | 1.73E-06 | 0.003951 |
| 5325 | PLAGL1 | 113.3530688 | 0.996035 | 0.209931 | 2.09E-06 | 0.004172 |
| 5592 | PRKG1 | 463.8615734 | 0.792318 | 0.172652 | 4.45E-06 | 0.007111 |
| 81551 | STMN4 | 407.1583395 | -0.97506 | 0.211744 | 4.13E-06 | 0.007111 |
| 2803 | GOLGA4 | 4826.145609 | 0.274702 | 0.060761 | 6.15E-06 | 0.008938 |
| 767 | CA8 | 198.8150717 | -0.9019 | 0.201271 | 7.43E-06 | 0.009888 |
| 1909 | EDNRA | 900.7642216 | 0.760681 | 0.17067 | 8.31E-06 | 0.010156 |
| 78997 | GDAP1L1 | 109.6847843 | -1.2734 | 0.286655 | 8.90E-06 | 0.010156 |
| 5175 | PECAM1 | 829.4028375 | 0.52938 | 0.12002 | 1.03E-05 | 0.010502 |
| 115548 | FCHO2 | 1475.189333 | 0.419033 | 0.0951 | 1.05E-05 | 0.010502 |
| 1290 | COL5A2 | 8963.612758 | 0.747587 | 0.170538 | 1.17E-05 | 0.010964 |
| 5205 | ATP8B1 | 252.8700371 | 0.809535 | 0.185212 | 1.24E-05 | 0.010983 |
| 7373 | COL14A1 | 860.8016235 | 0.945786 | 0.217792 | 1.41E-05 | 0.011838 |
| 9332 | CD163 | 12876.52315 | 1.308098 | 0.306163 | 1.93E-05 | 0.013013 |
| 9169 | SCAF11 | 4011.135905 | 0.300244 | 0.069986 | 1.79E-05 | 0.013013 |
| 875 | CBS | 961.7247316 | -0.49534 | 0.11545 | 1.78E-05 | 0.013013 |
| 93145 | OLFM2 | 808.9923179 | -0.58671 | 0.137404 | 1.96E-05 | 0.013013 |
| 441381 | LRRC24 | 33.97029124 | -0.90986 | 0.212358 | 1.83E-05 | 0.013013 |
| 3759 | KCNJ2 | 563.1353637 | 0.537025 | 0.126575 | 2.21E-05 | 0.013239 |
| 23216 | TBC1D1 | 1671.086313 | 0.291718 | 0.068805 | 2.24E-05 | 0.013239 |
| 345630 | FBLL1 | 33.27600773 | -1.23017 | 0.289728 | 2.18E-05 | 0.013239 |
| 26136 | TES | 368.73219 | 0.691061 | 0.16345 | 2.36E-05 | 0.013454 |
| 79187 | FSD1 | 176.8332367 | -0.60704 | 0.14422 | 2.56E-05 | 0.014123 |
| 4124 | MAN2A1 | 2722.209833 | 0.500494 | 0.119305 | 2.73E-05 | 0.014524 |
| 22925 | PLA2R1 | 289.4342541 | 0.796937 | 0.190545 | 2.88E-05 | 0.014864 |
| 1.01E+08 | DNM3OS | 135.1473547 | 1.177743 | 0.283332 | 3.23E-05 | 0.015651 |
| 84910 | TMEM87B | 790.3001554 | 0.373412 | 0.089841 | 3.23E-05 | 0.015651 |
| 5139 | PDE3A | 650.0727524 | 0.808889 | 0.195392 | 3.48E-05 | 0.016329 |
| 1378 | CR1 | 428.468695 | 1.721665 | 0.417006 | 3.65E-05 | 0.016655 |
| 84935 | MEDAG | 187.4729426 | 1.736515 | 0.422237 | 3.91E-05 | 0.016886 |
| 1601 | DAB2 | 1367.228764 | 0.89499 | 0.217557 | 3.89E-05 | 0.016886 |
| 3672 | ITGA1 | 3155.083673 | 0.892766 | 0.218458 | 4.38E-05 | 0.01705 |
| 55075 | UACA | 2801.043162 | 0.879256 | 0.215 | 4.32E-05 | 0.01705 |
| 4253 | MIA2 | 751.9145183 | 0.190349 | 0.046427 | 4.13E-05 | 0.01705 |
| 128312 | H2BU1 | 56.06297646 | -0.94549 | 0.231187 | 4.32E-05 | 0.01705 |
| 10184 | LHFPL2 | 2846.391379 | 0.668107 | 0.1639 | 4.58E-05 | 0.017402 |
| 57026 | PDXP | 301.3936541 | -0.66022 | 0.162233 | 4.71E-05 | 0.017496 |
| 1600 | DAB1 | 56.07243773 | -0.74573 | 0.183607 | 4.87E-05 | 0.017697 |
| 4060 | LUM | 1454.232745 | 1.585698 | 0.394083 | 5.73E-05 | 0.0182 |
| 728264 | CARMN | 714.9461116 | 1.138235 | 0.284377 | 6.27E-05 | 0.0182 |
| 1282 | COL4A1 | 25705.42138 | 0.927032 | 0.229689 | 5.44E-05 | 0.0182 |
| 1396 | CRIP1 | 139.1756583 | 0.882014 | 0.219741 | 5.97E-05 | 0.0182 |
| 3676 | ITGA4 | 1081.835497 | 0.598904 | 0.149273 | 6.02E-05 | 0.0182 |
| 29803 | REPIN1 | 985.3827012 | -0.28253 | 0.070259 | 5.79E-05 | 0.0182 |
| 728875 | NA | 71.47935093 | -0.54967 | 0.135915 | 5.25E-05 | 0.0182 |
| 1468 | SLC25A10 | 69.37581705 | -0.70006 | 0.174792 | 6.20E-05 | 0.0182 |
| 1152 | CKB | 2415.056651 | -0.88914 | 0.220577 | 5.55E-05 | 0.0182 |
| 494470 | RNF165 | 193.396958 | -0.94091 | 0.232927 | 5.36E-05 | 0.0182 |
| 11075 | STMN2 | 428.509595 | -1.54268 | 0.385229 | 6.21E-05 | 0.0182 |
| 9568 | GABBR2 | 317.4056257 | -1.42335 | 0.356861 | 6.65E-05 | 0.018965 |
| 4327 | MMP19 | 498.6702733 | 1.640407 | 0.416131 | 8.08E-05 | 0.020003 |
| 9509 | ADAMTS2 | 357.1242342 | 1.535422 | 0.389542 | 8.09E-05 | 0.020003 |
| 948 | CD36 | 401.8675628 | 1.213807 | 0.306493 | 7.49E-05 | 0.020003 |
| 9358 | ITGBL1 | 219.7353646 | 1.084708 | 0.274643 | 7.83E-05 | 0.020003 |
| 219623 | TMEM26 | 150.0464163 | 0.815724 | 0.206018 | 7.51E-05 | 0.020003 |
| 81792 | ADAMTS12 | 312.2400418 | 0.801566 | 0.202922 | 7.81E-05 | 0.020003 |
| 285203 | EOGT | 659.0670372 | 0.484049 | 0.12306 | 8.37E-05 | 0.020003 |
| 359845 | RFLNB | 399.7972356 | 0.467286 | 0.118812 | 8.39E-05 | 0.020003 |
| 7067 | THRA | 1744.471032 | -0.57394 | 0.145132 | 7.67E-05 | 0.020003 |
| 283248 | RCOR2 | 193.5926837 | -1.0851 | 0.275763 | 8.32E-05 | 0.020003 |
| 375704 | ENHO | 168.7048087 | -1.41831 | 0.360117 | 8.20E-05 | 0.020003 |
| 23317 | DNAJC13 | 3629.359927 | 0.181268 | 0.046274 | 8.96E-05 | 0.020776 |
| 90268 | OTULIN | 1024.722629 | 0.164369 | 0.041965 | 8.97E-05 | 0.020776 |
| 7855 | FZD5 | 453.0463433 | 0.510575 | 0.130528 | 9.17E-05 | 0.020922 |
| 140738 | TMEM37 | 115.1899765 | 0.92 | 0.23587 | 9.60E-05 | 0.021364 |
| 1496 | CTNNA2 | 2181.699365 | -0.4864 | 0.124725 | 9.63E-05 | 0.021364 |
| 1803 | DPP4 | 393.6808038 | 1.46781 | 0.377505 | 0.000101 | 0.022 |
| 51310 | SLC22A17 | 1001.60094 | -0.55445 | 0.142678 | 0.000102 | 0.022 |
| 57524 | CASKIN1 | 89.75823766 | -0.90651 | 0.233705 | 0.000105 | 0.02235 |
| 3559 | IL2RA | 322.7840705 | 1.407741 | 0.364006 | 0.00011 | 0.022532 |
| 1295 | COL8A1 | 785.7219972 | 0.829723 | 0.214528 | 0.00011 | 0.022532 |
| 7048 | TGFBR2 | 1865.961745 | 0.557067 | 0.143881 | 0.000108 | 0.022532 |
| 1303 | COL12A1 | 2159.00124 | 1.090289 | 0.283196 | 0.000118 | 0.023709 |
| 2335 | FN1 | 90865.15825 | 1.020001 | 0.265024 | 0.000119 | 0.023709 |
| 7476 | WNT7A | 60.23711675 | -0.64025 | 0.166501 | 0.00012 | 0.023743 |
| 23075 | SWAP70 | 1735.632773 | 0.261018 | 0.068001 | 0.000124 | 0.02412 |
| 4320 | MMP11 | 136.6725256 | 0.979662 | 0.255476 | 0.000126 | 0.024199 |
| 6362 | CCL18 | 165.8221877 | 3.206418 | 0.8398 | 0.000134 | 0.024614 |
| 3487 | IGFBP4 | 1330.361348 | 1.005899 | 0.264911 | 0.000146 | 0.024614 |
| 9060 | PAPSS2 | 534.3162319 | 0.717231 | 0.188686 | 0.000144 | 0.024614 |
| 4162 | MCAM | 3578.341451 | 0.554437 | 0.145774 | 0.000143 | 0.024614 |
| 2321 | FLT1 | 3181.634308 | 0.525557 | 0.137997 | 0.00014 | 0.024614 |
| 4094 | MAF | 1185.587343 | 0.470618 | 0.123125 | 0.000132 | 0.024614 |
| 9208 | LRRFIP1 | 1882.434969 | 0.334148 | 0.087816 | 0.000142 | 0.024614 |
| 5311 | PKD2 | 1730.840704 | 0.267787 | 0.070224 | 0.000137 | 0.024614 |
| 4597 | MVD | 198.203436 | -0.53868 | 0.141385 | 0.000139 | 0.024614 |
| 4915 | NTRK2 | 10188.32693 | -0.65318 | 0.171955 | 0.000146 | 0.024614 |
| 26232 | FBXO2 | 188.0271482 | -0.90256 | 0.237649 | 0.000146 | 0.024614 |
| 51617 | NSG2 | 330.3962473 | -1.48427 | 0.38894 | 0.000136 | 0.024614 |
| 4625 | MYH7 | 116.0433084 | -0.7667 | 0.202208 | 0.00015 | 0.024902 |
| 1278 | COL1A2 | 10955.85651 | 1.338037 | 0.354165 | 0.000158 | 0.025246 |
| 54829 | ASPN | 244.8656243 | 1.23206 | 0.325763 | 0.000156 | 0.025246 |
| 57125 | PLXDC1 | 1016.990503 | 0.824465 | 0.218261 | 0.000158 | 0.025246 |
| 154141 | MBOAT1 | 132.9346344 | 0.808017 | 0.21415 | 0.000161 | 0.025246 |
| 30061 | SLC40A1 | 1744.7296 | 0.43417 | 0.115026 | 0.00016 | 0.025246 |
| 138311 | DIPK1B | 300.7735757 | -0.46717 | 0.123724 | 0.000159 | 0.025246 |
| 6781 | STC1 | 751.7672591 | 1.197282 | 0.317934 | 0.000166 | 0.025498 |
| 54796 | BNC2 | 387.8907464 | 0.787387 | 0.208994 | 0.000165 | 0.025498 |
| 4026 | LPP | 7269.585995 | 0.49995 | 0.132952 | 0.00017 | 0.025568 |
| 6538 | SLC6A11 | 255.5257214 | -1.00746 | 0.267829 | 0.000169 | 0.025568 |
| 8338 | H2AC20 | 610.1125553 | -0.58486 | 0.155735 | 0.000173 | 0.025828 |
| 2444 | FRK | 45.02494024 | 0.807139 | 0.215587 | 0.000181 | 0.02634 |
| 2200 | FBN1 | 3836.740463 | 0.689311 | 0.184011 | 0.00018 | 0.02634 |
| 10924 | SMPDL3A | 308.1054271 | 0.670435 | 0.179163 | 0.000183 | 0.02634 |
| 9697 | TRAM2 | 777.0399409 | 0.634394 | 0.169563 | 0.000183 | 0.02634 |
| 8654 | PDE5A | 931.6264987 | 0.644951 | 0.172872 | 0.000191 | 0.02706 |
| 51560 | RAB6B | 1252.662182 | -0.53887 | 0.144465 | 0.000191 | 0.02706 |
| 55803 | ADAP2 | 953.2199966 | 0.61838 | 0.165884 | 0.000193 | 0.027066 |
| 203522 | INTS6L | 432.7831097 | 0.268365 | 0.072046 | 0.000195 | 0.027139 |
| 5551 | PRF1 | 194.6917989 | 1.233593 | 0.331646 | 0.0002 | 0.027242 |
| 4643 | MYO1E | 1517.086364 | 0.580884 | 0.156161 | 0.000199 | 0.027242 |
| 3912 | LAMB1 | 6558.519009 | 0.952069 | 0.256399 | 0.000205 | 0.027469 |
| 27 | ABL2 | 2714.176905 | 0.308347 | 0.08303 | 0.000204 | 0.027469 |
| 11326 | VSIG4 | 3914.818203 | 1.06168 | 0.286626 | 0.000212 | 0.02801 |
| 10522 | DEAF1 | 529.1343407 | -0.28902 | 0.078005 | 0.000211 | 0.02801 |
| 144402 | CPNE8 | 334.4890902 | 0.631622 | 0.170661 | 0.000215 | 0.028114 |
| 23175 | LPIN1 | 1576.370062 | -0.27185 | 0.073812 | 0.000231 | 0.029701 |
| 1826 | DSCAM | 836.6862168 | -0.89297 | 0.242412 | 0.00023 | 0.029701 |
| 121457 | IKBIP | 710.8423048 | 0.456273 | 0.124168 | 0.000238 | 0.03044 |
| 5300 | PIN1 | 505.7290614 | -0.2893 | 0.078845 | 0.000243 | 0.030466 |
| 23580 | CDC42EP4 | 2054.20444 | -0.5124 | 0.139757 | 0.000246 | 0.030466 |
| 844 | CASQ1 | 102.851126 | -0.62236 | 0.169543 | 0.000242 | 0.030466 |
| 55964 | SEPTIN3 | 919.5956037 | -0.87965 | 0.239917 | 0.000246 | 0.030466 |
| 677828 | SNORA47 | 176.1461107 | -0.49527 | 0.135231 | 0.00025 | 0.030707 |
| 221395 | ADGRF5 | 1868.792441 | 0.592567 | 0.162029 | 0.000255 | 0.030861 |
| 59342 | SCPEP1 | 1196.295538 | 0.478 | 0.130639 | 0.000253 | 0.030861 |
| 57089 | ENTPD7 | 391.9679173 | 0.532667 | 0.145848 | 0.00026 | 0.031226 |
| 8685 | MARCO | 535.2883966 | 2.576484 | 0.70937 | 0.000281 | 0.031385 |
| 114904 | C1QTNF6 | 189.8862482 | 0.889486 | 0.245128 | 0.000285 | 0.031385 |
| 1368 | CPM | 886.7084237 | 0.739726 | 0.203536 | 0.000279 | 0.031385 |
| 11214 | AKAP13 | 6303.945295 | 0.338091 | 0.092877 | 0.000272 | 0.031385 |
| 10921 | RNPS1 | 1012.036306 | -0.16959 | 0.046521 | 0.000267 | 0.031385 |
| 51222 | ZNF219 | 186.2433482 | -0.36032 | 0.099032 | 0.000274 | 0.031385 |
| 79007 | DBNDD1 | 248.8072732 | -0.39024 | 0.107414 | 0.00028 | 0.031385 |
| 3757 | KCNH2 | 289.7502657 | -0.5773 | 0.158908 | 0.00028 | 0.031385 |
| 23542 | MAPK8IP2 | 253.6324494 | -0.69246 | 0.190819 | 0.000285 | 0.031385 |
| 9363 | RAB33A | 76.79579553 | -0.86168 | 0.236206 | 0.000264 | 0.031385 |
| 162494 | RHBDL3 | 397.04649 | -0.95185 | 0.261879 | 0.000278 | 0.031385 |
| 10690 | FUT9 | 1111.737231 | -1.03623 | 0.28554 | 0.000285 | 0.031385 |
| 51696 | HECA | 1206.177572 | 0.254234 | 0.070139 | 0.000289 | 0.031558 |
| 374875 | HSD11B1L | 155.8397076 | -0.35897 | 0.09906 | 0.00029 | 0.031558 |
| 84892 | POMGNT2 | 534.3183284 | -0.39647 | 0.109508 | 0.000294 | 0.031737 |
| 5799 | PTPRN2 | 575.3596982 | -0.46404 | 0.129048 | 0.000323 | 0.034659 |
| 132720 | FAM241A | 76.4253114 | 0.88035 | 0.245329 | 0.000333 | 0.034731 |
| 22795 | NID2 | 874.5732207 | 0.822026 | 0.228902 | 0.000329 | 0.034731 |
| 9695 | EDEM1 | 1378.143463 | 0.459338 | 0.12798 | 0.000332 | 0.034731 |
| 155185 | AMZ1 | 90.55429743 | -0.70038 | 0.195173 | 0.000333 | 0.034731 |
| 4017 | LOXL2 | 1457.428712 | 0.995069 | 0.277577 | 0.000337 | 0.034986 |
| 51393 | TRPV2 | 314.0000926 | 0.668317 | 0.186517 | 0.000339 | 0.034987 |
| 1374 | CPT1A | 1281.279583 | 0.492052 | 0.137443 | 0.000344 | 0.035179 |
| 23040 | MYT1L | 219.6461449 | -1.58061 | 0.441958 | 0.000348 | 0.035445 |
| 7402 | UTRN | 8581.245903 | 0.315511 | 0.088689 | 0.000374 | 0.037852 |
| 1284 | COL4A2 | 11792.98347 | 0.860112 | 0.242032 | 0.00038 | 0.038159 |
| 2261 | FGFR3 | 971.5925916 | -0.73788 | 0.207961 | 0.000388 | 0.038734 |
| 10052 | GJC1 | 1509.788124 | 0.453422 | 0.127883 | 0.000392 | 0.038853 |
| 57084 | SLC17A6 | 62.45771606 | -2.08441 | 0.588143 | 0.000394 | 0.038853 |
| 1E+08 | WHAMMP1 | 332.1194823 | 0.488077 | 0.138238 | 0.000414 | 0.040369 |
| 4713 | NDUFB7 | 564.652786 | -0.42705 | 0.120918 | 0.000413 | 0.040369 |
| 4312 | MMP1 | 333.0243407 | 2.357464 | 0.669073 | 0.000426 | 0.041023 |
| 10912 | GADD45G | 122.9919302 | -1.2734 | 0.361557 | 0.000428 | 0.041023 |
| 114794 | ELFN2 | 170.5098342 | -1.57895 | 0.448354 | 0.000429 | 0.041023 |
| 4642 | MYO1D | 416.4996328 | 0.805727 | 0.23039 | 0.00047 | 0.043232 |
| 1075 | CTSC | 3371.493764 | 0.737876 | 0.210894 | 0.000467 | 0.043232 |
| 55614 | KIF16B | 690.9023115 | 0.338261 | 0.096565 | 0.00046 | 0.043232 |
| 5978 | REST | 1979.939898 | 0.336081 | 0.096128 | 0.000472 | 0.043232 |
| 23607 | CD2AP | 1148.566558 | 0.310308 | 0.088542 | 0.000457 | 0.043232 |
| 130612 | TMEM198 | 185.5749796 | -0.45957 | 0.131483 | 0.000474 | 0.043232 |
| 56967 | C14orf132 | 2304.611287 | -0.55489 | 0.158338 | 0.000458 | 0.043232 |
| 53826 | FXYD6 | 1795.861564 | -0.776 | 0.222009 | 0.000473 | 0.043232 |
| 8600 | TNFSF11 | 17.21627915 | 1.563785 | 0.448565 | 0.00049 | 0.043657 |
| 3101 | HK3 | 469.1411728 | 1.060946 | 0.303903 | 0.000481 | 0.043657 |
| 133584 | EGFLAM | 198.9539668 | 1.056084 | 0.303028 | 0.000492 | 0.043657 |
| 8038 | ADAM12 | 1172.340227 | 0.752182 | 0.2158 | 0.000491 | 0.043657 |
| 23086 | EXPH5 | 272.076768 | -0.593 | 0.170044 | 0.000488 | 0.043657 |
| 5069 | PAPPA | 487.7788881 | 1.45492 | 0.417961 | 0.0005 | 0.043837 |
| 1306 | COL15A1 | 500.8326701 | 1.293265 | 0.371434 | 0.000498 | 0.043837 |
| 4323 | MMP14 | 4320.572043 | 0.607981 | 0.174775 | 0.000504 | 0.043837 |
| 3688 | ITGB1 | 11064.35875 | 0.540154 | 0.155421 | 0.00051 | 0.043837 |
| 29965 | CDIP1 | 575.7181599 | -0.3303 | 0.095043 | 0.00051 | 0.043837 |
| 55228 | PNMA8A | 1059.662381 | -0.52364 | 0.150668 | 0.00051 | 0.043837 |
| 23640 | HSPBP1 | 306.095839 | -0.28365 | 0.081887 | 0.000532 | 0.04548 |
| 79812 | MMRN2 | 390.28342 | 0.577056 | 0.166768 | 0.00054 | 0.045616 |
| 83539 | CHST9 | 431.0490011 | -0.60842 | 0.175791 | 0.000538 | 0.045616 |
| 10882 | C1QL1 | 281.9994608 | -0.775 | 0.224379 | 0.000552 | 0.04644 |
| 9890 | PLPPR4 | 779.0493132 | -0.6626 | 0.192343 | 0.000571 | 0.047567 |
| 2066 | ERBB4 | 707.39527 | -0.98605 | 0.286254 | 0.000572 | 0.047567 |
| 1824 | DSC2 | 431.3027964 | 0.806546 | 0.234836 | 0.000594 | 0.049132 |
| 284716 | RIMKLA | 240.5064459 | -0.5165 | 0.150504 | 0.0006 | 0.049369 |
| 9211 | LGI1 | 126.2127386 | -0.86347 | 0.251749 | 0.000604 | 0.049465 |
| 55702 | YJU2 | 270.6322614 | -0.26324 | 0.076798 | 0.000609 | 0.049608 |

***Supplementary Table 2: Differential expression results****. Differential gene expression between ‘stiff’ and ‘soft’ biopsies in 8 glioblastoma patients (22 biopsies). Only genes with an adjusted p-value (Benjamini and Hochberg method) below 0.05 are shown. Genes with higher expression in ‘stiff’ biopsies have log2foldchange>0, genes with higher expression in ‘soft’ biopsies have log2foldchange<0.*
